# Molecular insights into mitoepigenetic stress response signaling in age-associated cardiovascular disease risk

**DOI:** 10.1101/2024.03.12.24304065

**Authors:** Nikita Soni, Prasan Kaur, Vikas Gurjar, Arpit Bhargava, Rajnarayan Tiwari, Vinay Singh Raghuwanshi, Rupesh K. Srivastava, Pradyumna Kumar Mishra

## Abstract

Cardiovascular diseases (CVD) represent a major challenge for the elderly population and continue to be among the leading causes of mortality worldwide. The integrity of mitochondria in peripheral lymphocytes can indicate age-related degenerative disorders, revealing the balance between bioenergetics, inflammation, and senescence. The present study aims to investigate the impact of aging on mitochondrial-mediated epigenetic stress response mechanisms in both young and elderly individuals. Mitochondrial oxidative DNA damage, repair response, copy number, methylation, fusion, fission genes, electron transport chain enzyme complex activities, pro-inflammatory cytokine levels, and associated epigenetic mechanisms were studied. The results demonstrate that the elderly group experienced more significant disruption in the expression of mitochondrial genes, leading to impaired mitochondrial DNA (mtDNA) integrity intricately linked to mitochondrial-nuclear crosstalk, increasing their susceptibility to CVD. The findings highlight mitochondria’s crucial role in cellular dynamics, influencing stress responses and epigenetic mechanisms through fusion and fission cycles. Overall, these insights provide valuable information about the complex molecular processes linking mitochondrial dysfunction to age-related cardiovascular risks, paving the way for further exploration and potential interventions in aging-related health outcomes.

**Graphical Abstract:** Figure showing the molecular mechanisms underlying the mitoepigenetic stress response signaling pathways in age-associated cardiovascular diseases.

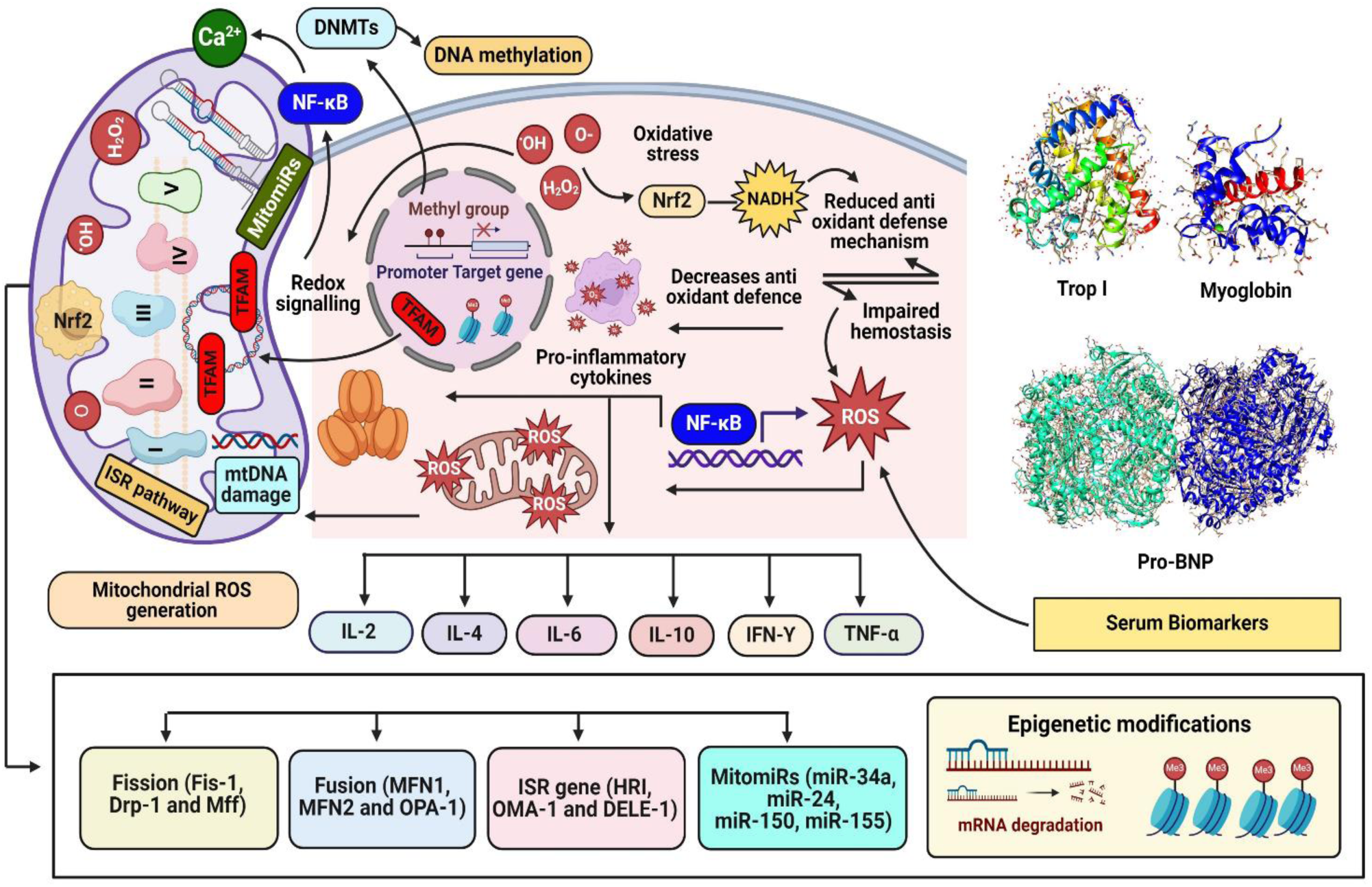

## 1. Introduction

Cardiovascular diseases (CVDs), affecting 40% of the elderly population, stand out as predominant age-related ailments and a leading cause of global morbidity and mortality [1]. Due to its status as a key regulator of cellular metabolism, research on the function of mitochondria in the development and course of CVDs has been extensive [2,3]. Mitochondria, renowned as cellular powerhouses undergo various functional and morphological changes that results in the onset and progression of CVDs. These dynamic organelles undergo shape and subcellular distribution changes, controlled by mitochondrial fusion and fission cycles, in response to external factors [3]. Numerous studies have suggested that fusion supports interconnected mitochondria biogenesis and fission results in mitochondrial fragments, thereby affecting the different mitochondrial functions. Such perturbations within mitochondrial machinery disturb the regulatory cascade of reactive oxygen species (ROS), thereby, resulting in excessive and uncontrolled release of these reactive molecules. In addition, a growing body of evidence suggests that uncontrolled ROS could further lead to mtDNA heteroplasmy, impaired calcium homeostasis, and deficient mitophagy and subsequently increases the oxidative mtDNA damage [4, 5]. Importantly, the damage induced by ROS to mtDNA manifests as compromised production of enzymes critical for oxidative phosphorylation, further intensifying mitochondrial dysfunction and promoting increased cellular senescence [6]. In general, to exclude such impairment, mitochondrial repair mechanisms such as base excision repair (BER), single-strand break repair (SSBR), mismatch repair (MMR), or double-strand break repair (DSBR) pathways are involved, but significantly BER pathway is triggered to eliminate this damage [7,8]. Moreover, studies also indicate the mitochondrial ROS can cause the activation of inflammatory mediators such as transcription factor NF-κB [9,10]. The activation of NF-kB might further stimulate the release of pro-inflammatory cytokines, such as TNF-α and IL-6. Furthermore, the release of mtDNA into the cytoplasm, which can happen because of ruptured micronuclei, can activate the MAPK and cyclic GMP–AMP synthase (cGAS) pathways, which in turn can activate the inflammatory pathways [11]. It has been shown that increased mtDNA in plasma source exosomes in chronic heart disease causes an inflammatory response through the TLR9-NF-κB pathway [12]. Some studies have also reported that binding of TNF-α to its receptors induces downstream phosphorylation of p47phox (phox: phagocyte oxidase) and facilitates nicotinamide adenine dinucleotide phosphate hydrogen (NADPH) oxidase activity, which further results in mitochondrial dysfunction and mtDNA damage [13]. In response to such disruptions, an adaptive response pathway commonly known as integrated stress response (ISR) is activated [14]. Predominantly, this mechanism is crucial for balanced cellular homeostasis by coordinating with mitochondrial unfolded protein response in an eIF2α dependent pathway [15]. Earlier studies have reported that aging triggers a pro-survival signalling mechanism inside the cell and can induce mito-epigenetic modifications either directly or indirectly. These unfavourable epigenetic changes cause alterations in the mitochondrial DNA methylation facilitated by DNA methyltransferases and expression profile of microRNAs (miRNAs) [16]. Our previous studies have documented the role of disturbed mitochondrial machinery in modifications in the mtDNA methylation and miRNAs expression. As the detailed mechanisms of age associated mito-epigenetic stress response processes are not well known, we herein aimed to understand the potential mechanistic insights within different age cohorts. Our objective was to comprehend mitochondrial dysfunction and assess various parameters, including mtDNA damage levels, inflammation, integrated stress responses, methylation, and mitochondrial complexes activity in peripheral blood lymphocytes as promising biomarkers for cardiovascular risk.

## 2. Material & methods

### 2.1 Reagents and kits

All the reagents used were of the highest available grade. Lymphocytes were isolated using HiSep™ (HiMedia Laboratories, Mumbai, MH, India) and 1× PBS from Cell Signaling Technology, Danvers, MA, USA. Whole-cell DNA was obtained using the PureLink Genomic DNA Mini Kit from Invitrogen by Life Technology-Thermo Fisher Scientific, USA. Total RNA isolation was carried out using the TRIzol® Reagent from Life Technologies - Thermo Fischer Scientific, Waltham, MA, USA. CellROX® Deep Red Flow Cytometry assay kit from Thermo Fischer Scientific, Waltham, MA, USA was used for the Oxidative stress measurement. New England Biolabs’ Luna® Universal One-Step RT-qPCR Kit (Ipswich, MA, USA) was used to quantify mitochondrial fission and fusion, signalling, and survival genes using Real time-qPCR. The Luna® Universal qPCR Master Mix from New England Biolabs, (Ipswich, MA, USA), was used to measure mtDNA methylation and DNMT1 levels, and the relative expression of mitomiRs and their target genes as well. To evaluate the expression of mitochondrial genes using conventional PCR, Taq 2X Master Mix purchased from New England Biolabs, Ipswich, MA, USA was used. BamHI restriction enzymes from Takara Bio Inc., Shiga, Japan, and EpiJET Bisulfite Conversion Kit from Thermo Fischer Scientific, Waltham, MA, USA, were used to analyze mtDNA methylation. Poly(A) Tailing kit and PureLink miRNA isolation kit from Invitrogen-Thermo Fischer Scientific, Waltham, MA, USA, were used for mitomiRs profiling. SeaKem® LE Agarose from Lonza, Basel, Switzerland was used for gel electrophoresis, along with 50× TAE and 6X gel loading buffer from HiMedia Laboratories, Mumbai, MH, India; SYBR® Safe DNA gel stain from Invitrogen-Thermo Fischer Scientific, Waltham, MA, USA; 100 bp DNA Ladder from HiMedia Laboratories Pvt. Ltd. Mumbai, India, and 50 bp DNA Ladder from Promega, Corporation, Madison, WI, USA were used.

### 2.2. Study design

In this study, comparison between the alterations in mitochondrial inflammation and integrated stress response in two different age groups, group I (n= 154; individuals aged 18-38 years) and group II (n= 105; individuals aged 39-65 years and older), using a lymphocyte model was done. The study was approved by the Institutional Human Ethics Committee (IHEC, ICMR-National Institute for Research in Environmental Health, Bhopal, India) and was carried out with funding from the Department of Health Research (DHR), Ministry of Health and Family Welfare (MoHFW), Government of India, with strict adherence to sample collection guidelines. A total of 259 blood samples were collected for the study from various cities in Madhya Pradesh, India, and were divided into two groups. Lymphocytes were chosen as the cellular system to conduct the studies being an attractive *in vitro* model system that accurately mimics host immunological interactions. Blood samples were collected, and lymphocytes separated using HiSep™ by density gradient centrifugation at 500Xg for 30 minutes. To explore mitochondrial dysfunction, we extracted whole DNA, RNA, and protein. The expression levels of mitochondrial fission and fusion genes Drp1, Fis1, Mff, MFN1, MFN2, and OPA1, as well as the mitochondrial genes, MT-ATPase6, and MT-ATPase8, MT-CO1, MT-ND6 were assessed to understand mitochondrial dynamics and functioning. Furthermore, we analysed the expression levels of genes associated with the ISR (DELE1, HRI, and OMA1) and the repair process (POLG, OGG, and APE). We also evaluated mtDNA methylation in the D-loop, 12S, cytochrome B, and 16S rRNA regions, as well as mitochondrial DNA methyltransferase1 (DNMT1, DNMT3a, and DNMT3b) and H3 alterations as epigenetic markers. We also looked at the expression patterns of mitochondrial miRNAs (mitomiRs), including mitomiR24, mitomiR34a, mitomiR150, and mitomiR155.The inflammatory markers i.e., IL-6, IL-4, IL-2, IL-10, INF-γ and TNF-α was also analysed to check the age associated inflammation. Clinical biomarkers including Trp I, Mb and NT-pro-BNP were also analysed to test the prevalent age associated cardiac diseases risk.

### 2.3. Analysis of oxidative DNA damage

To assess oxidative damage, the occurrence of oxidized purine bases (commonly deoxyguanosine) was examined using the formamidopyrimidine glycosylase (FPG) digestion method. 100 ng of DNA sample was incubated for 1 h at 37^◦^C in 10µL of reaction mixture containing 1 unit of FPG enzyme, 10 mM Bis Tris Propane-HCl, 10 mM MgCl2, 1 mM DTT, and 0.1 mg/mL bovine serum albumin. Oxidative damage was analyzed through quantitative RT-qPCR (Insta Q-96™, HiMedia Laboratories, Mumbai, MH, India). Oxidative injury in mtDNA was detected through increased oxidised purine nucleotides. The results were presented as the fold change was calculated by using the ddCt method, which represents the difference between the Ct values in β-actin and mitogenes treated with FPG enzyme (Ct-T) and samples not treated with FPG enzyme (Ct-UT). A greater ΔCt value indicates augmented oxidative stress in the mitochondria [14].

### 2.4. Evaluation of mitochondrial repair genes

The mitochondrial repair genes’ expression levels were determined through quantitative analysis of OGG-1, APE, and POLG genes, following a method previously reported. In brief, total RNA isolation from peripheral lymphocytes was performed through the standard phase separation method. The isolated RNA was then pooled based on different age groups and quantified. The prepared pooled samples were then mixed with the reaction mixture comprising specific primers and the Luna® Universal One-Step RT-qPCR Master Mix followed by the amplification on Insta Q-96™ (HiMedia Laboratories, Mumbai, MH, India). The GAPDH was used as an internal control. Ct values were recorded, and fold change was calculated by using ddCt method [17].

### 2.5. Assessment of survival mechanism (integrated stress response)

To evaluate survival mechanisms, RT-qPCR (Insta Q-96™, HiMedia Laboratories, Mumbai, MH, India) was used to compare expression levels of OMA1, DELE1, and HRI genes implicated in integrated stress across age groups. After isolation, an approximate quantity of RNA for 20 ng was amplified by using Luna® Universal One-Step RT-qPCR Kit and respective primers. GAPDH was used as an internal control. After the completion of the PCR cycles, Ct values were obtained, and fold change was calculated by using the ddCt method [18].

### 2.6. Evaluation of inflammatory cytokine level

To examine the inflammatory response. IL-6, TNF-α, and IFN-γ levels were measured using Human GENLISATM ELISA kits from KRISHGEN BioSystems, US. All the instructions from the protocol manual were strictly adhered and the readings were recorded by using Spark® multimode microplate reader (TECAN, Seestrasse 103, Männedorf, Switzerland) [19].

### 2.7. Analysis of mitochondrial fission and fusion genes

RT-qPCR (Insta Q-96™, HiMedia Laboratories, Mumbai, MH, India) was used to analyze the relative expression of mitochondrial fission genes Drp1, Fis1, and Mff, as well as fusion genes MFN 1, MFN2, and OPA1. Total RNA was extracted and amplified using the previously described method. GAPDH served as an internal control. Following amplification, Ct readings were obtained, and the fold change was calculated using the ddCt approach [20].

### 2.8. TFAM’s binding to promoter regions

This experiment was performed using a set of TFAM-specific primers and the previously disclosed RT-qPCR method [12]. TET belongs to the methylcytosine dioxygenase family of enzymes that demethylate DNA. TET1, TET2, and TET3 were evaluated using the RT-qPCR approach described elsewhere, with a specific set of primers [21].

### 2.9. Estimation of mt-DNA methylation

To examine the aging-associated epigenetic mtDNA methylation profile, DNA was extracted and measured. After quantification, 1000 ng DNA was mixed with 15 µl 10X Buffer, 3 µl restriction endonuclease BamH1, and nuclease-free water to make up a volume of up to 75 µl. The resulting mixture was incubated for four hours at 37°C before being treated with bisulfite. To convert bisulfite, mix 15 µl of BamH1-treated DNA with 125 µl of bisulfite mix. Incubate at 95°C for 5 minutes, 65°C for 60 minutes, 95°C for 5 minutes, 65°C for 60 minutes, 95°C for 5 minutes, and 65°C for 90 minutes. Desulphonation and cleanup were then performed. The bisulphite-converted DNA was amplified with Excel TaqTM 2X Q-PCR Master Mix (SYBR, ROX) Smobio master mix by RT-qPCR. A PCR reaction with 50 ng bisulfite-converted primer DNA in a 25 µl volume was amplified for 35 cycles. GAPDH served as an internal control. GAPDH served as an internal control. Ct readings were acquired following the scheduled PCR cycles, and the fold change was calculated using the ddCt approach [14].

### 2.10. Relative gene expression analysis of epigenetic modifiers (DNA methyltransferase)

The expression levels of epigenetic modifiers DNMT1, DNMT3a, and DNMT3b were determined using a Real Time-PCR method (Insta Q96TM, HiMedia Laboratories, Mumbai, MH, India). The analysis was done by employing a set of DNMT1, DNMT3a, and DNMT3b specific primers and following the RT-qPCR procedure as described above (section 2.4) [19].

### 2.11. Analysis of mitochondrial genes

ATPase6, ATPase8, COX1, and ND6 genes genes were evaluated to analyze the expression of mitochondrial genes. After quantifying RNA samples, an approximate 20 ng of RNA was amplified using the Luna® Universal One-Step RT-qPCR Kit and adding respective primers on RT-qPCR. GAPDH served as an internal control. After completing PCR cycles, Ct values were acquired, and fold change was computed using the ddCt method [22].

### 2.12. Measurement of mitochondrial electron chain complexes activity

MitoCheck Complex Activity Assay kits from Cayman, Ann Arbour, MI, USA (700930 for Complex I, 700940 for Complex II, 700950 for Complex III, 700990 for Complex IV, and 701000 for Complex V) were used to investigate age-related changes in mitochondrial activity. The isolated protein sample was obtained and processed following the assay methodology. The readings were recorded using Spark® multimode microplate reader (TECAN, Seestrasse 103, Männedorf, Switzerland) [23].

### 2.13. miRNA expression profiling

As abnormal miRNA expression in old age is linked to the onset and progression of several noncommunicable diseases, the expression profiling of miR34a, miR150, miR155, and miR24 was carried out using the methods outlined below. In brief, extracted miRNA was poly-adenylated, cDNA synthesised, and amplification using appropriate primers on an Insta Q96TM real-time PCR (HiMedia Laboratories, Mumbai, MH, India). We employed U6 as an internal control. After the scheduled PCR cycles, Ct values were recorded, and dCt values were calculated by subtracting the Ct value of the internal control from the Ct value of the miRNAs [12].

### 2.14. Characterization of histones

ELISA was used to determine post-translational epigenetic modifications of histone H3 protein, including methylation (mono-, di-, and tri-methylation of lysine residues at 4, 9, 27, 36, and 79th positions), acetylation (at 9, 14, and 18th positions of lysine), and phosphorylation (at 10 and 28th positions of serine residues). Briefly, antibody buffer and histone extracts were added to the antibody-coated wells and incubated for 90 minutes. Following incubation, the wells were cleaned, and HRP-conjugated antibody added. Following color development, a stop solution was used to terminate the reaction and readings were recorded at 450 nm using Spark® multimode microplate reader [12].

### 2.15. Detection of CVD biomarkers

100 ul of biotinylated NT-proBNP antibody working solution was added to each well. The plate was sealed, incubated at 37°C for 60 minutes, and rinsed four times with diluted Wash Buffer (1X). 100 ul Streptavidin:HRP conjugate working solution was pipetted into all wells and mixed, followed by 100 ul TMB. Substrate was added to all the wells and incubated for 10 minutes at 37°C. Positive wells should turn bluish, and 100 ul of stop solution was added to each well. The wells should change from blue to yellow in colour. Absorbance was measured at 450 nm on a microplate within 10-15 minutes after adding the Stop solution.

### 2.16. Statistics

All experiments were reported as mean ± standard error from sample sets. All data sets were examined with GraphPad Prism 9. We used a two-way ANOVA with multiple comparisons to determine significance among both groups. Furthermore, in this study, we employed the R programming language (R Studio) to analyze correlations among age, mtDNA/nDNA ratio, mtDNA methylation, and proBNP levels. Our focus was on uncovering insights with potential implications for biological and clinical contexts. The dataset that served as the basis for our further study was created using the outcomes of our thorough investigation. Using Pearson correlation coefficients calculated with R’s cor() function, we explored the strength and direction of relationships between these key variables. The illustration of the correlations was given via the correlation matrix and scatterplot matrix that were produced. While acknowledging the limitations and assumptions inherent in correlation analyses, our study adhered to rigorous scientific standards.

## 3. Results

### 3.1 Demographic data of subjects

The demographic data for subjects of both groups (289) enrolled in the study are listed in Supplementary Table 1. Compared with healthy subject, the body mass index (BMI), waist circumference, blood pressure, cholesterol, and troponin I levels were higher in group II as compared to group I (Supplementary Table 1).

### 3.2 Oxidative DNA damage in cells

The analysis of 8-oxo-dG, a modified nucleotide base used to detect oxidative DNA damage, revealed its time-dependent accumulation in the cells. The observations also indicated that mtDNA damage, which is age-related endothelial deterioration in arteries is significantly correlated with an increase in mitochondrial reactive ROS production. Group I cell had the highest 8-oxo-dG levels 2.76±0.8 ng/mL, whereas group II has shown levels of 4.07±0.7 ng/mL (Figure 1).

**Figure 1:**
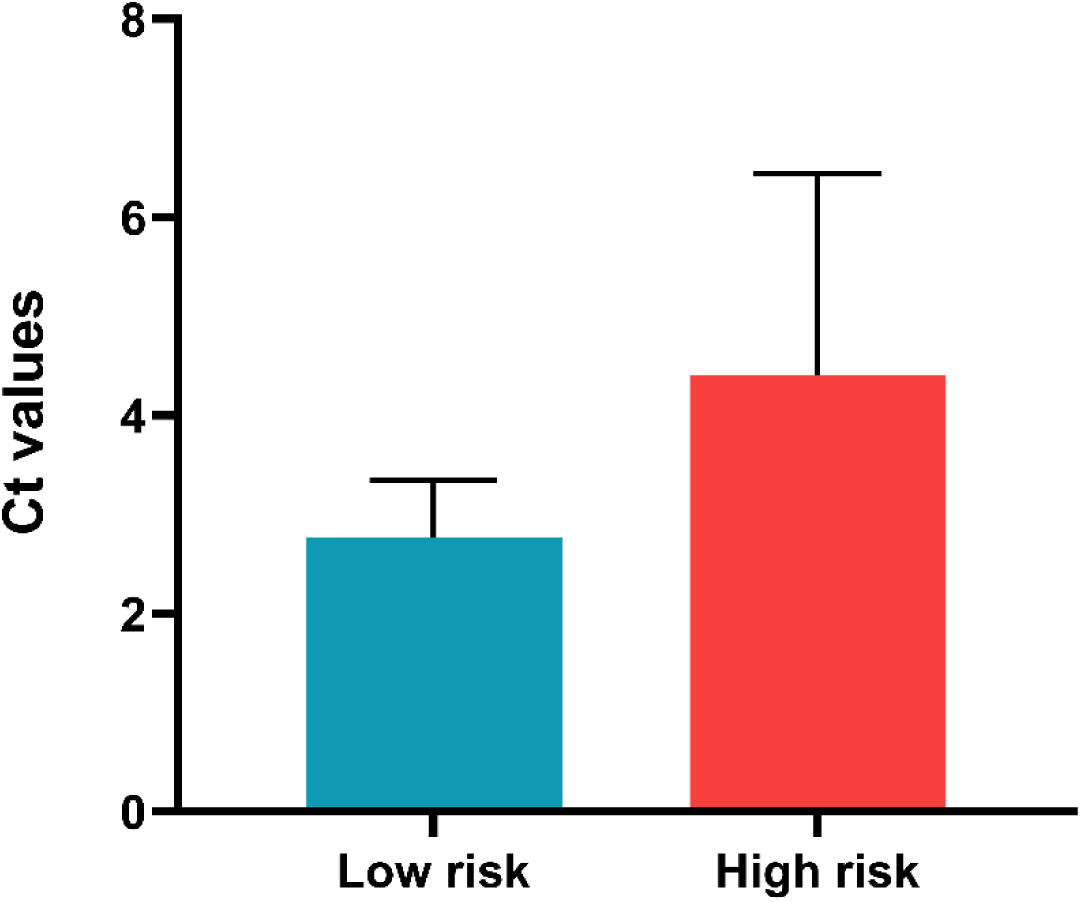
Mitochondrial oxidative DNA damage. Graph showing the levels of oxidative DNA damage among group I and in group II samples estimated by FPG digestion. The levels of oxidative mtDNA damage are shown as del Ct, which is the difference between threshold cycle (Ct) values between enzyme - treated and enzyme - non-treated samples (Ct-T and Ct-N, respectively).

### 3.3 Activation of the mitochondrial repair cascade

As part of its protective mechanism, mtDNA damage triggers a mitochondrial base-excision repair response that eliminates the accretion of mutations in the form of small and bulky DNA adducts induced by oxidative damage, primarily affecting the OGG1, APE, and POLG genes. The study discovered that the OGG1, APE, and POLG genes were more highly expressed in older age groups, with fold changes of 1.05±0.65, 0.99±0.16, and 2.07±0.81, respectively (Figure 2).

**Figure 2:**
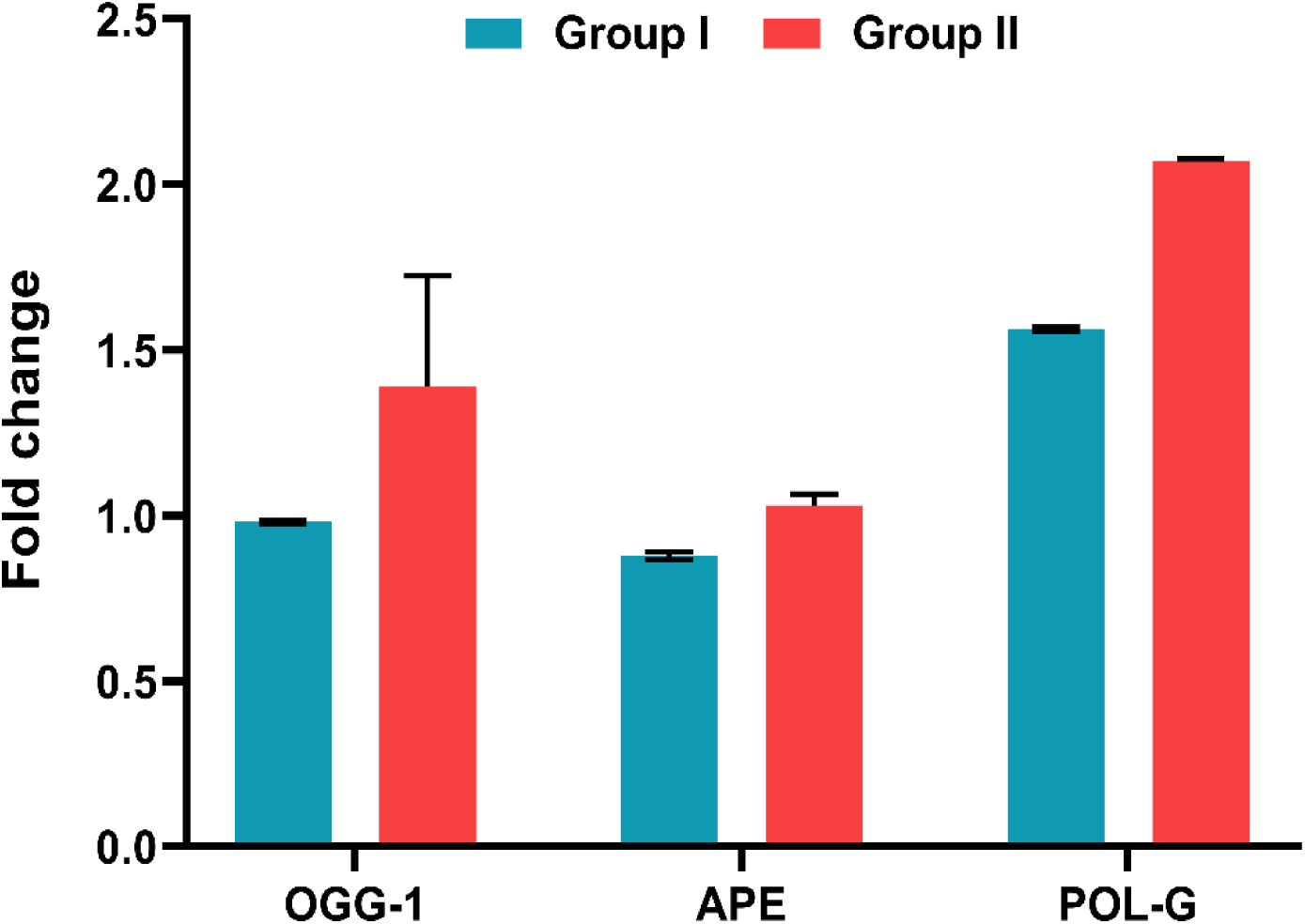
Mitochondrial DNA repair cascade. Graphical representation of the fold expression observed in different mitochondrial DNA repair genes. The fold change of group I and in group II was calculated as 2^-ΔΔCt by identifying dCt as the difference of internal control from their respective control and test values.

### 3.4 Stimulated mitochondrial integrated stress response

Aging is responsible for declination in bio-energetic and lacking in the antioxidant defense mechanism, which radially increases the generation of ROS in the cells, and oxidatively stressed cells may trigger a pro-survival signalling pathway like ISR to keep the cellular homeostasis intact. The results showed upregulated expression of the ISR genes (OMA-1, DELE-1, and HRI) in the older age group as compared to the younger age group. The observed fold change for OMA-1, DELE-1, and HRI in the older age group was 2.23±0.64, 1.69±0.45 and 1.30±56 respectively (Figure 3).

**Figure 3:**
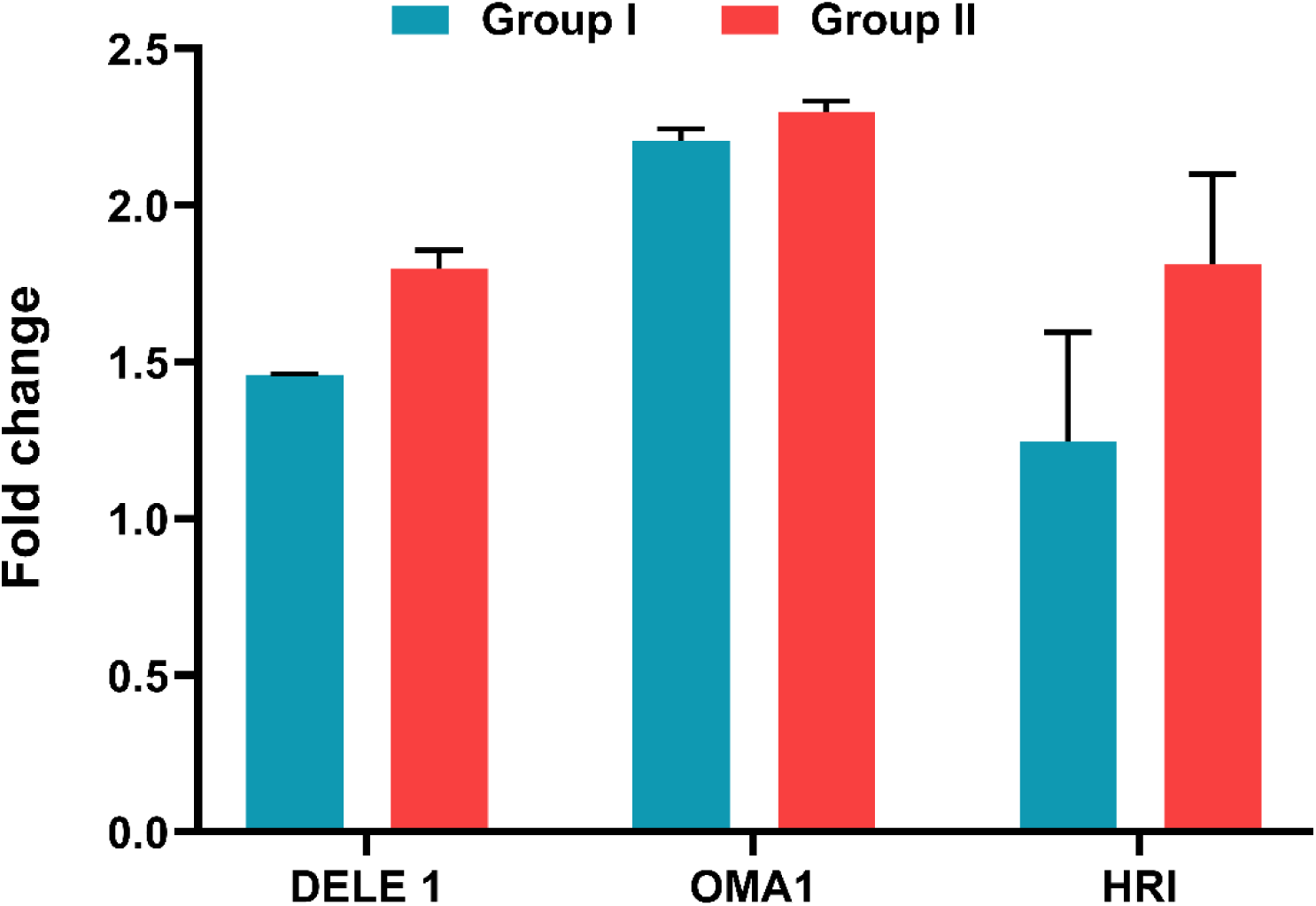
Integrated stress response. A barplot representing fold change in the expression levels of ISR genes OMA1, DELE1, and HRI genes in group I and in group II. The fold change was calculated as 2^-ΔΔCt by identifying dCt as the difference of internal control from their respective control and test values.

### 3.5 Irregular inflammatory responses

Pro-inflammatory responses were activated in both groups, with older individuals exhibiting higher levels of IL-6, TNF-α, and IFN-γ compared to younger risk groups. In the older age group, IL-6, TNF-α, and IFN-γ levels were 182.5 ± 5.15 pg/mL, 22.01 ± 1.50 pg/mL, and 55.29 ± 5.19 pg/mL, respectively. In the younger age group, the values were 11.97 ± 1.002 pg/mL, 10.57 ± 0.83 pg/mL, and 10.66 + 0.62 pg/mL (Figure 4).

**Figure 4:**
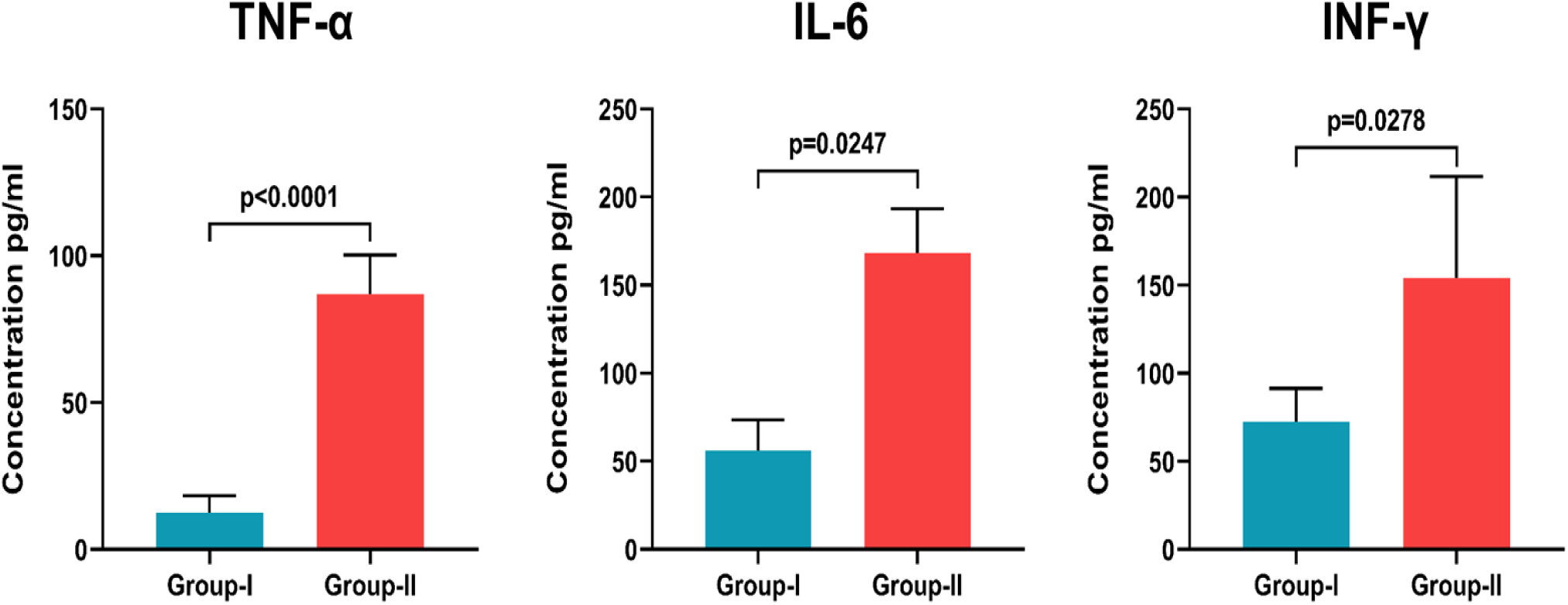
Pro-inflammatory cytokine levels. Graph showing the levels of pro-inflammatory cytokines in cell exposed to different concentration of group I and in group II. The values are expressed as mean ± SE and p ≤ 0.05 was considered significant.

### 3.6. Alterations in mitochondrial biogenesis (fusion and fission)

The coordinated linked cycles of fission and fusion that underpin mitochondrial biogenesis change depending on the cell’s metabolic requirements. The quantities of Drp1, Fis1, MFF (fission genes), OPA1, MFN1 and MFN2 (fusion genes) detected in this investigation varied significantly across older persons. Drp1, Fis1, MFF, MFN1, MFN2, and OPA1 in the elder age group displayed a mean relative fold change of 3.01±0.36, 4.08±0.86, 1.37±0.16, 3.32±0.56, 0.53±0.38, and 3.71±74, respectively. The outcomes also suggested that the genome and mitochondrial dynamics might be unstable (Figure 5).

**Figure 5:**
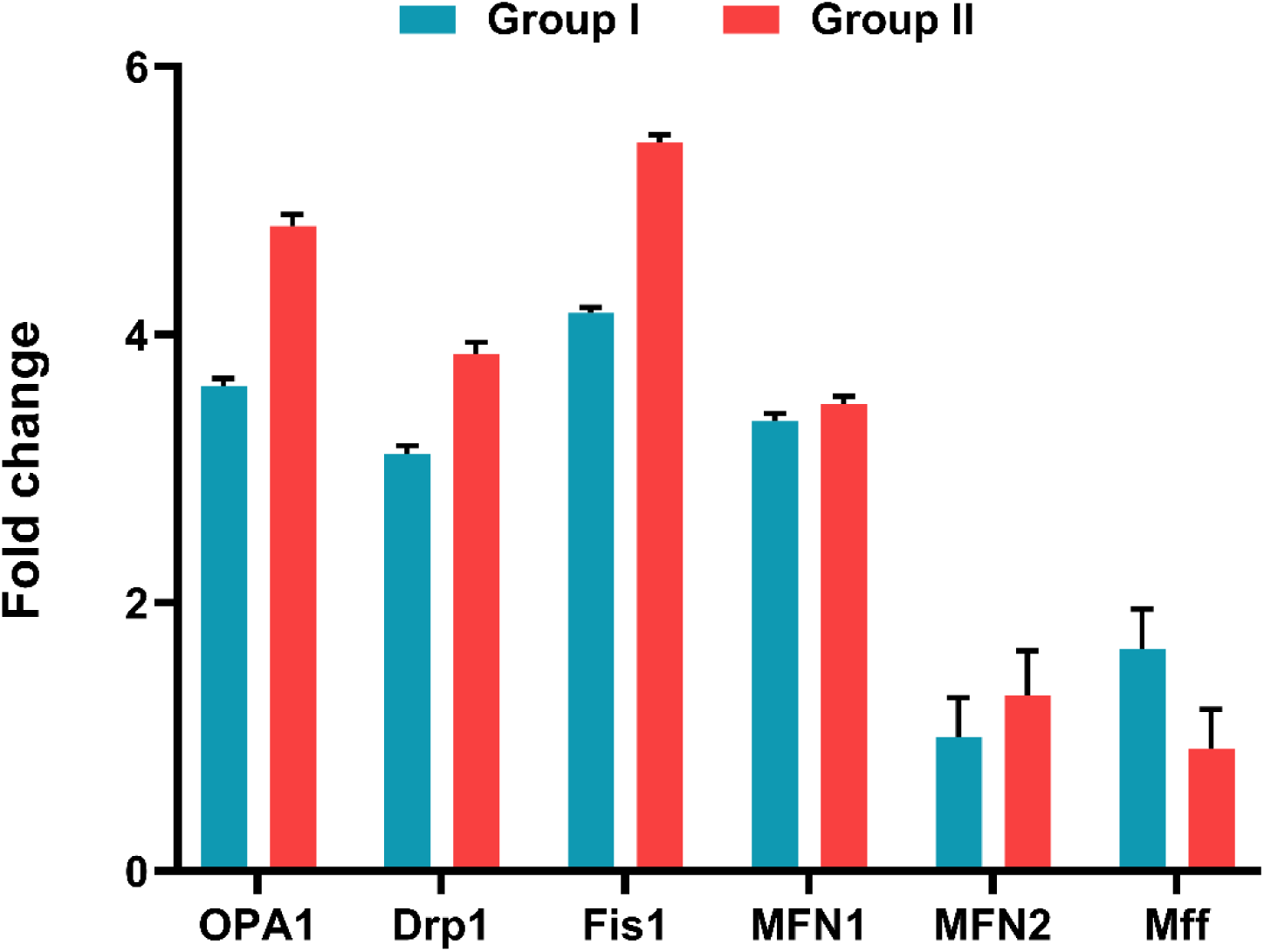
Mitochondrial fission and fusion genes. Graph showing the change in fold expression in different mitochondrial fission and fusion genes in group I and in group II. The fold change was calculated as 2^-ΔΔCt by identifying (dCt) as the difference of internal control from their respective control and test values.

### 3.7. Activation of the mitochondrial transcription factor and ten-eleven translocation

The mitochondrial transcription factor TFAM is an important regulator of mtDNA replication and transcription. In the presence of changed stimuli, the TFAM plays a vital function in the orientation and expression of mitochondrial genes, governing the proper response. The gene expression was significantly higher in the older age group, with a fold change of 6.74±0.49. Similarly, TET1, TET2, and TET3 revealed higher gene expression in the older age group compared to the younger group. The measured fold change was 4.31±0.23, 4.09±0.65, and 3.95±0.23 (Figure 6).

**Figure 6:**
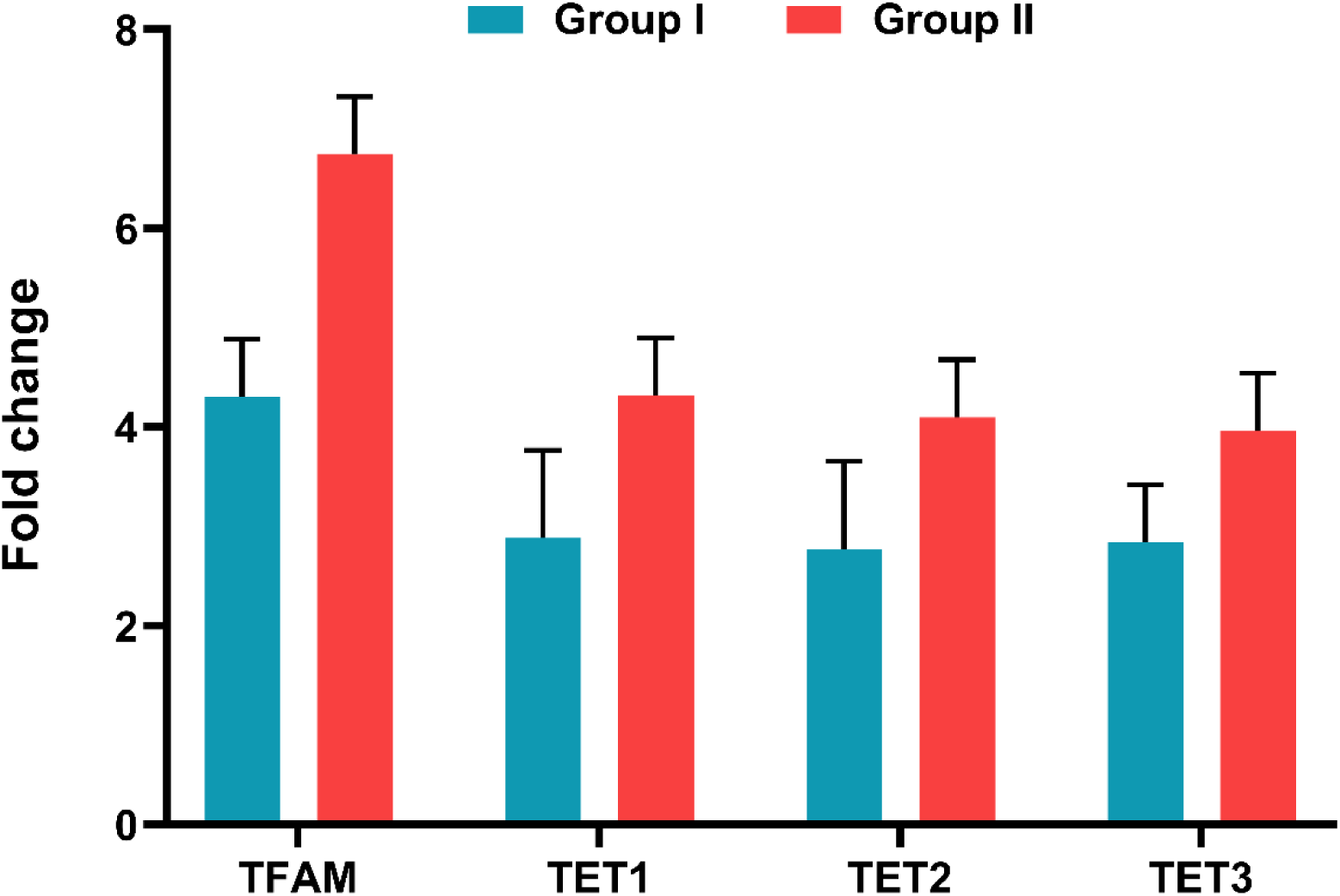
Mitochondrial TFAM and TET. A bar plot representing the fold change in the expression of mitochondrial TFAM and TET1, 2 and 3 in group I and in group II. The fold change was calculated as 2^− ΔΔCt by identifying dCt as the difference of internal control from their respective control and test values.

### 3.8. Disturbed mtDNA methylation machinery

The study also analyzed at the methylation status of five mtDNA loci (D-loop1, D-loop2, 12S + TF, CYTB, and 16S) in high-risk and low-risk groups. The findings suggested that age and other environmental factors cause considerable changes to the mitochondrial methylation profile. The examination of mitochondrial areas, including the displacement loop, 12S +TF, cytochrome B, and 16S rRNA, revealed methylation differences between Group II and Group I. In group II, the methylation levels for D-loop1, D-loop2, 12S + TF, CYT B, and 16S were 6.49±0.86, 3.68±0.49, 2.60±0.13, 1.95±0.49, and 1.02±0.58, respectively. In group I, the values were 4.56±1.11, 0.72±0.64, 1.33±0.17, 1.10±0.63, and 3.13±0.74 (Figure 7).

**Figure 7:**
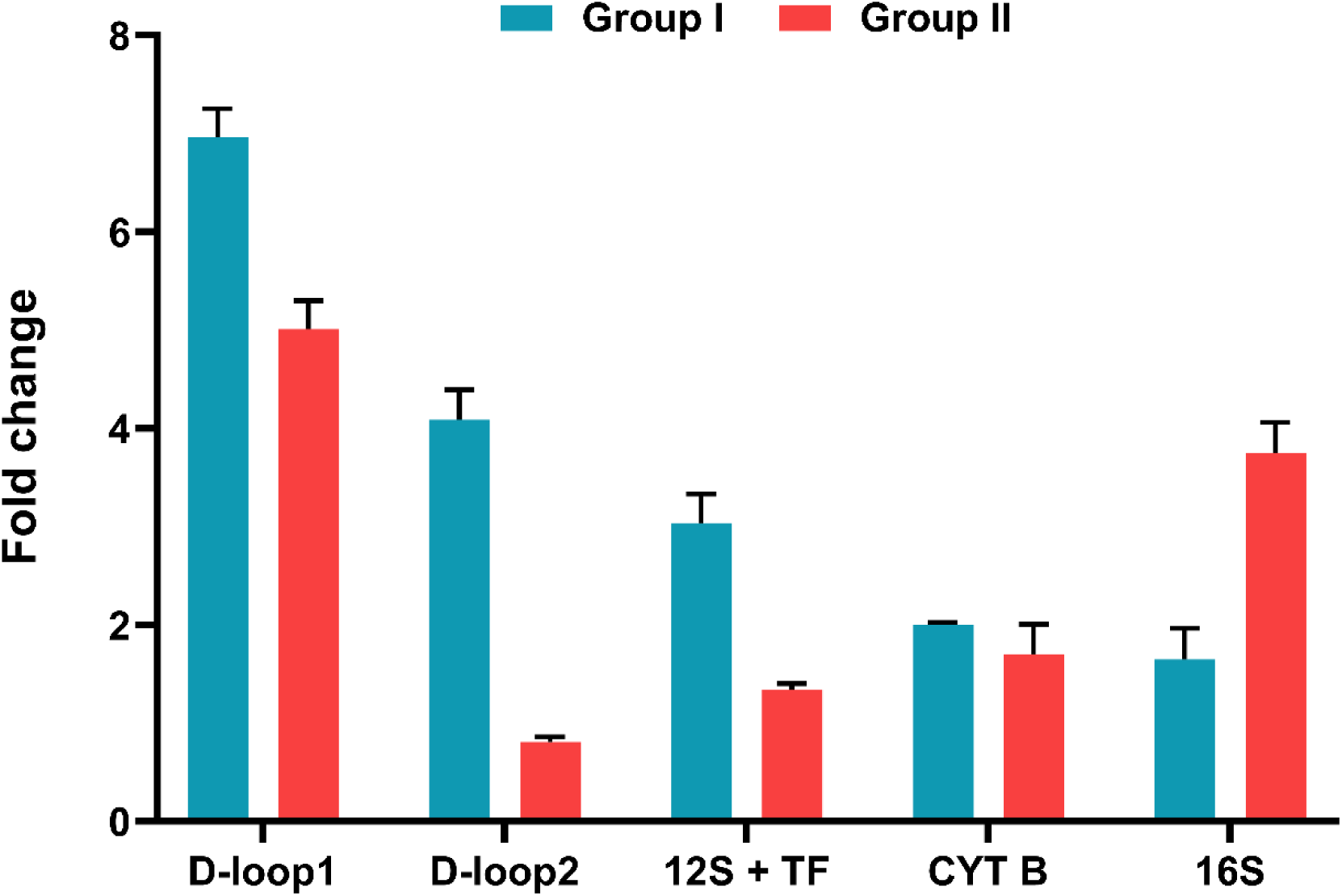
**Disturbed mtDNA methylation machinery**. Graph showing the status of methylation (fold change) within D-loop 1, D-loop 2, 12S-TF, CYTB and 16S regions of mtDNA among group I and in group II. The fold change was calculated as 2^-ΔΔCt by identifying dCt as the difference of internal control from their respective control and test values.

### 3.9. DNA methyltransferase gene expression

Mitochondrial biogenesis is linked with ongoing mitochondrial-epigenetic processes, particularly DNMT-regulated mtDNA methylation. This dynamic epigenetic process involves DNMT enzymes, including DNMT1, DNMT3a, and DNMT3b, which catalyze the addition of methyl groups to DNA. A higher level of damage recruits the most DNMTs in the process, resulting in increased methylation. The modifications in the expression of DNMT-1, 3a, and 3b in group II was observed to be 8.17±0.12, 6.41±0.09, and 5.99±0.13, while group I levels were 6.25±0.96, 4.06±0.64, and 3.98±0.76 respectively (Figure 8).

**Figure 8:**
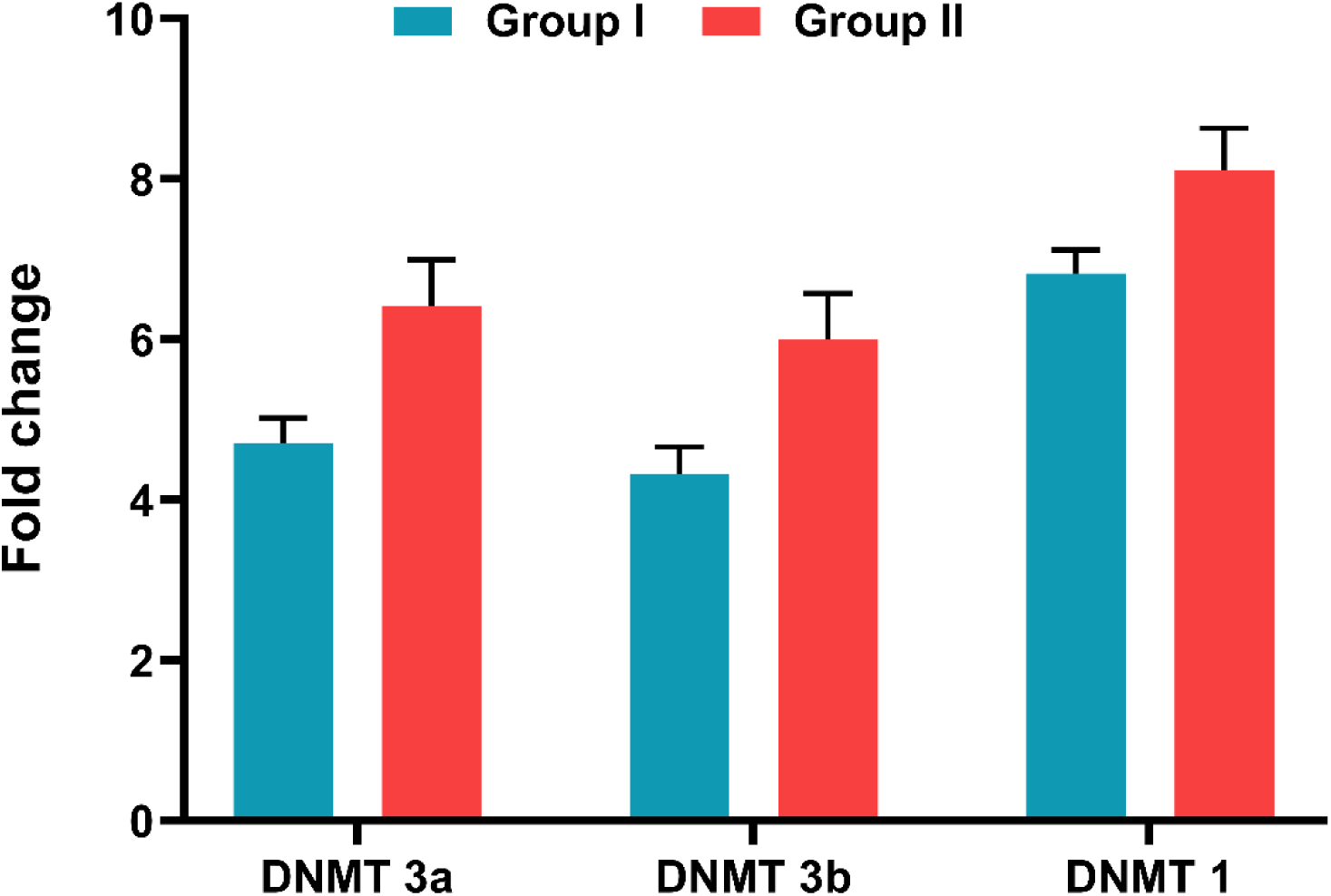
Expression of DNA methyltransferase. Graph showing the real-time quantification of DNMT1, 3a and 3b in the pooled samples of group I and in group II. The fold change was calculated as 2^-ΔΔCt by identifying dCt as the difference of internal control from their respective control and test values.

### 3.10. Aging-related mitochondrial dysfunction

Aging-related changes to the mitochondrial genome may lead to dysregulation of essential mitochondrial machinery that controls genes including ND-6, Cyt-OX1, ATPase-6, and ATPase-8. These genes serve as components of mitochondrial respiratory chain complexes and are necessary for proper mitochondrial activity. In comparison to young age group I, older age group II showed increased expression of mitochondrial genes. The fold change for ATPase6, ATPase8, COX1, and ND6 was 0.0059±0.009, 0.0074±0.006, 0.0092±0.010, and 0.0025±0.003 respectively. These findings revealed that older people have worse mitochondrial function (Figure 9).

**Figure 9:**
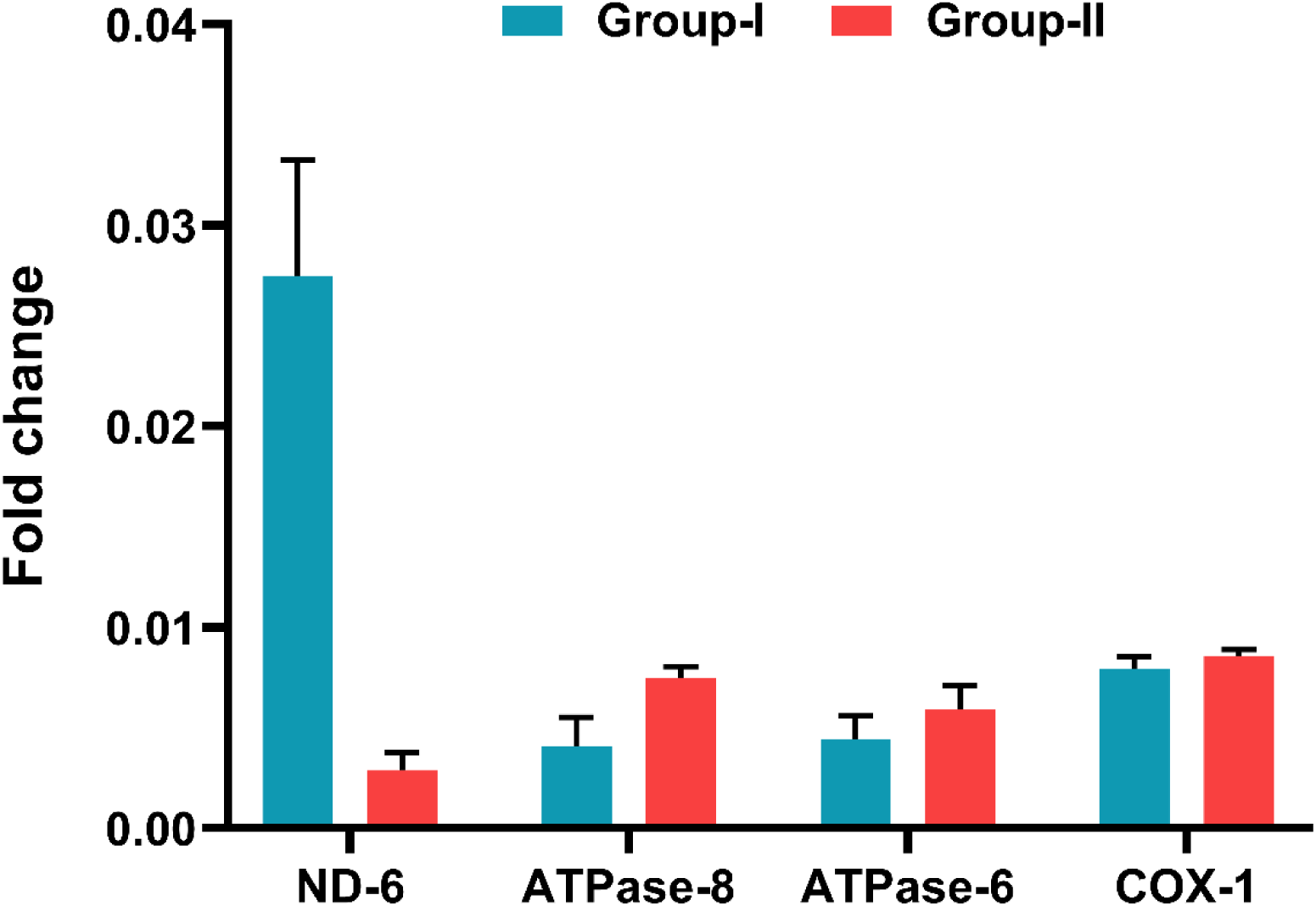
Mitochondrial genes. Graphical representation of the fold change in the expression of mitochondrial genes in group I and group II. The fold change was calculated as 2^-ΔΔCt by identifying dCt as the difference of internal control from their respective control and test values.

### 3.11. Mitochondrial respiratory chain enzymes

Compared to the low-risk group, Group I had higher levels of mitochondrial gene expression. This raises the possibility of abnormalities in the functioning of the mitochondria. This observation was supported by significant differences in the activity of the mitochondrial electron chain complexes (Complex I, II, III, IV, and V). The results revealed differences in the activity of these complexes in group II compared to group I. Group II’s mean specific activities for complexes I, II, III, IV, and V were 47.05±2.11, 38.16±11.9, 27.63±5.16, 73.9±3.07, and 67.65±12.17, and 42.79±2.85, 34.59±2.90, 21.44±4.36, 65.65±4.45, and 58.53±18.2 in Group I, respectively (Figure 10).

**Figure 10:**
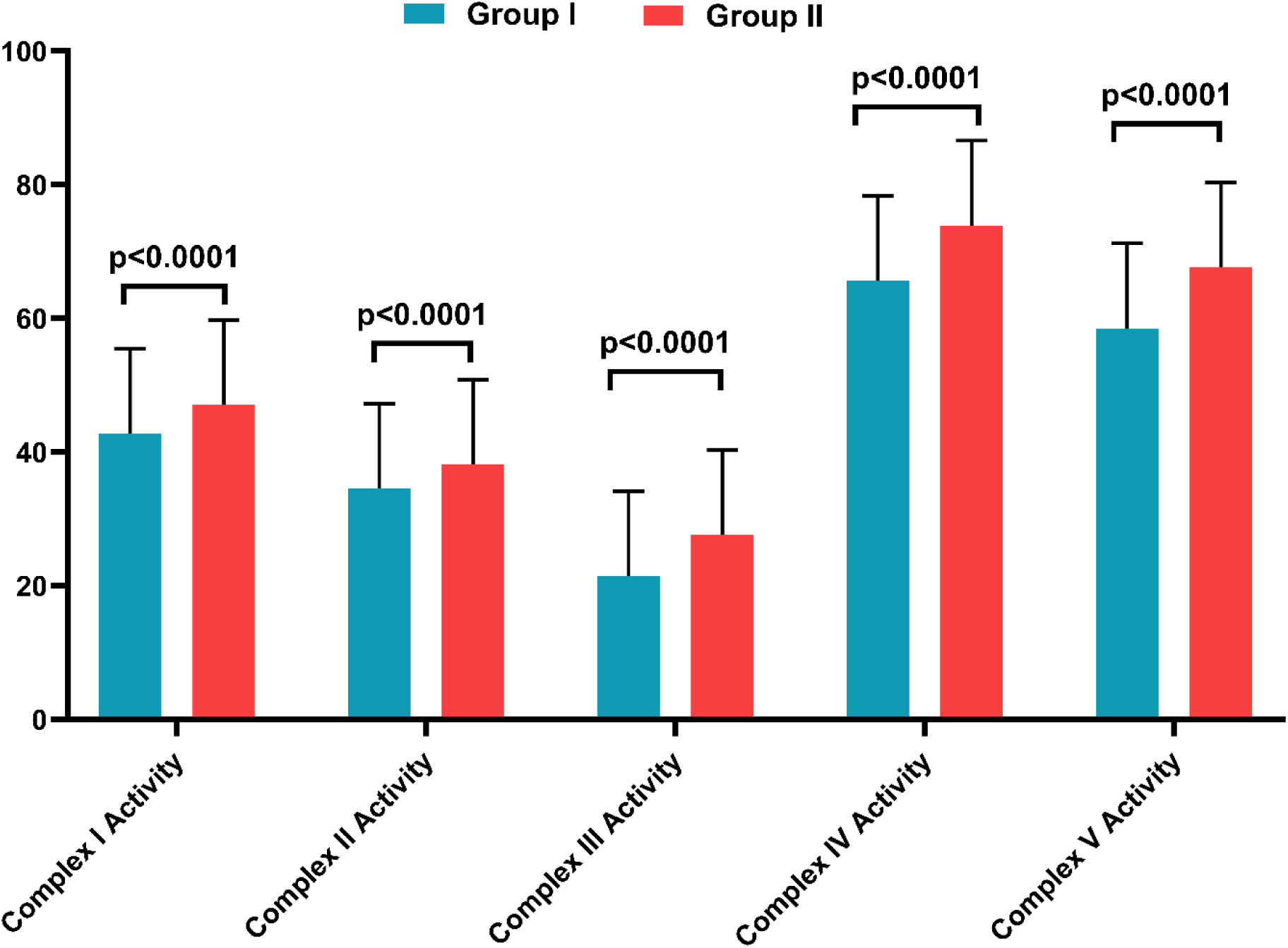
Mitochondrial complex activity. A barplot showing the mean activity of mitochondrial electron chain complexes (complex I, II, III, IV and V) group I and in group II. The values are expressed as mean ± SE and p ≤ 0.05 was considered significant.

### 3.12. Changes in the expression profile of mitomiR

miRNAs are small, highly conserved RNAs that regulate gene expression. MitomiRs are also responsible for regulating epigenetic alterations in mtDNA. The increased expression of the TET 1, TET 2, TET 3, and TFAM genes promotes mtDNA methylation while decreasing transcription machinery at transcription sites. This process affects the expression levels of microRNAs and long non-coding RNAs. The results revealed differences in the expression of mitomiRs in group I. Group II had greater fold change values for mitomiR24, mitomiR150, mitomiR34a, and mitomiR155 compared to group I (3.29±0.88, 1.09±0.16, 2.66±0.83, and 2.21±0.90, respectively) (Figure 11).

**Figure 11:**
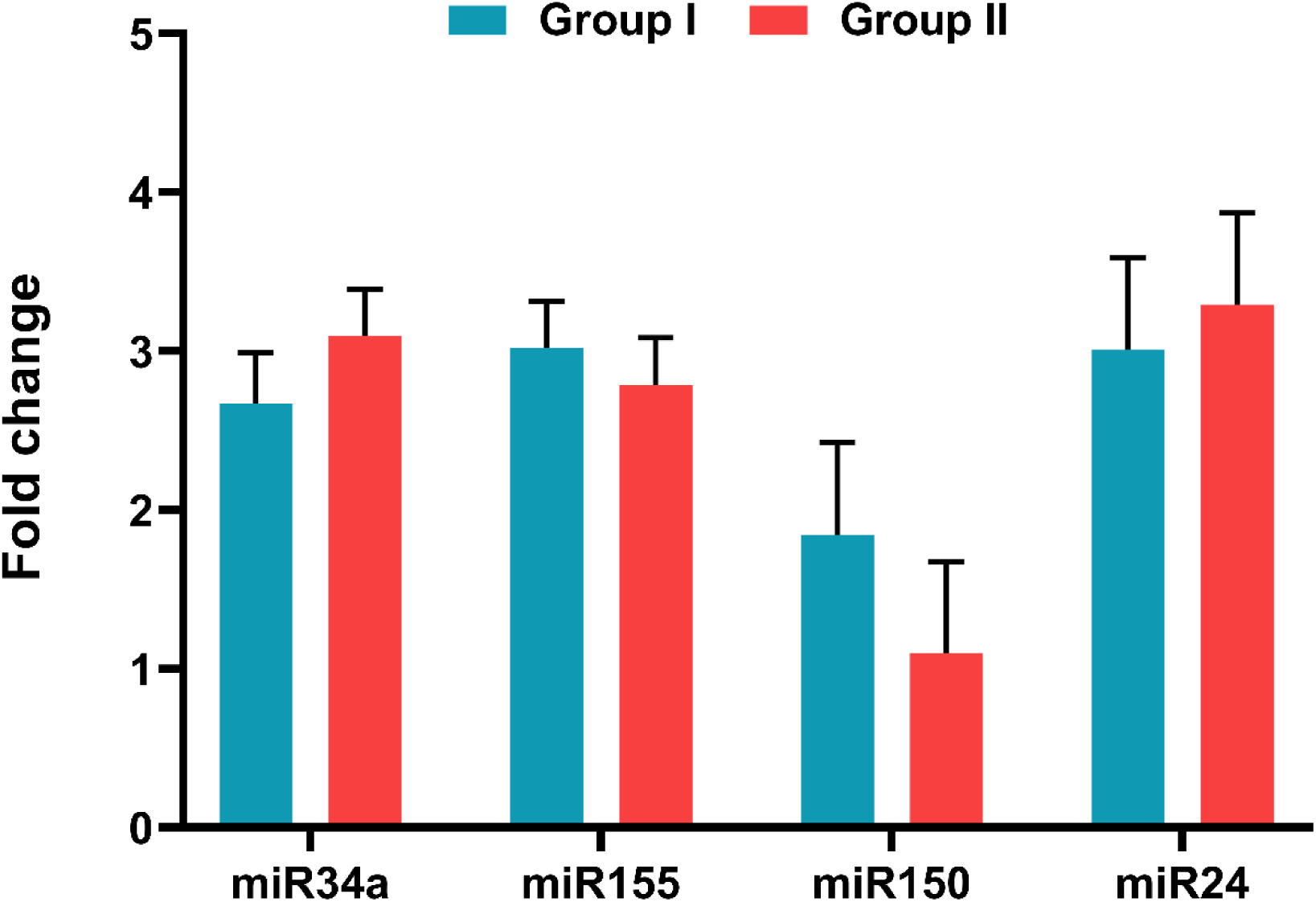
mitomiR profiling. The graph showing the expression profile of mitomiRs among group I and in group II. The fold change was calculated as 2^-ΔΔCt by identifying dCt as the difference of internal control from their respective control and test values.

### 3.13. Histone H3 modifications

Histone modifications have a crucial role in regulating the activation or inhibition of several proteins that are engaged in essential physiological processes, including cell cycle regulation, proliferation of cells, and apoptosis. For example, H3K4 and H3K36 methylation is associated with active transcription, whereas H3K9, H3K27, and H4K20 methylation are associated with transcriptional repression. In the current study, the results demonstrated variations in histone methylation (H3K4me1, H3K4me3, H3K9me2, H3K79me3), acetylation (H3K9ac) and phosphorylation patterns (H3Ser10P) among in elderly (group II), when compared to healthy individuals (group I) whereas deregulation in methylation levels (H3K4me2, H3K9me1, H3K9me3, H3K27, H3K36, H3K79me2), acetylation (H3K14ac, H3K18ac) and phosphorylation patterns (H3Ser10P) among group I as compared to group II (Figure 12).

**Figure 12:**
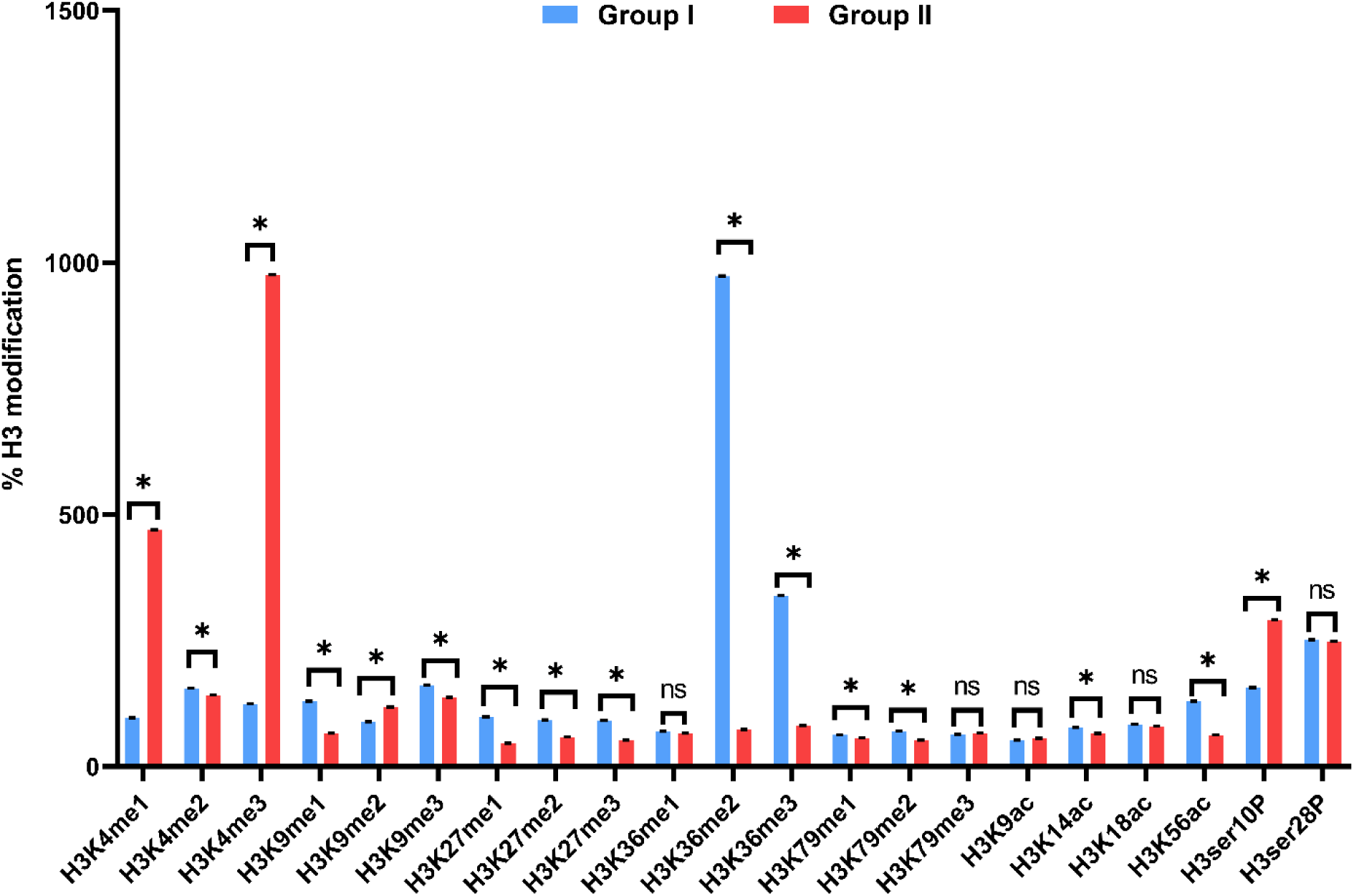
Histone H3 modifications. The graph showing the analysis of 21 histone H3 modifications in group I and group II. The values are expressed as mean ± SE. NS represents non-significant, while the asterisk denotes p ≤ 0.05.

### 3.14. Detection of CVD Biomarkers

In group II, cardiac Trp I has a concentration of 120.76±10.06 compared to the younger age group of 68.58±8.34, Mb in group I with 7.390±1.37 and group II with 21.61±1.202, and Pro-BNP concentrations of 113.61±37.074 compared to the risk group of 141.1402± 47.66484, indicating a high risk of cardiac biomarkers in the elderly (Figure 13).

**Figure 13:**
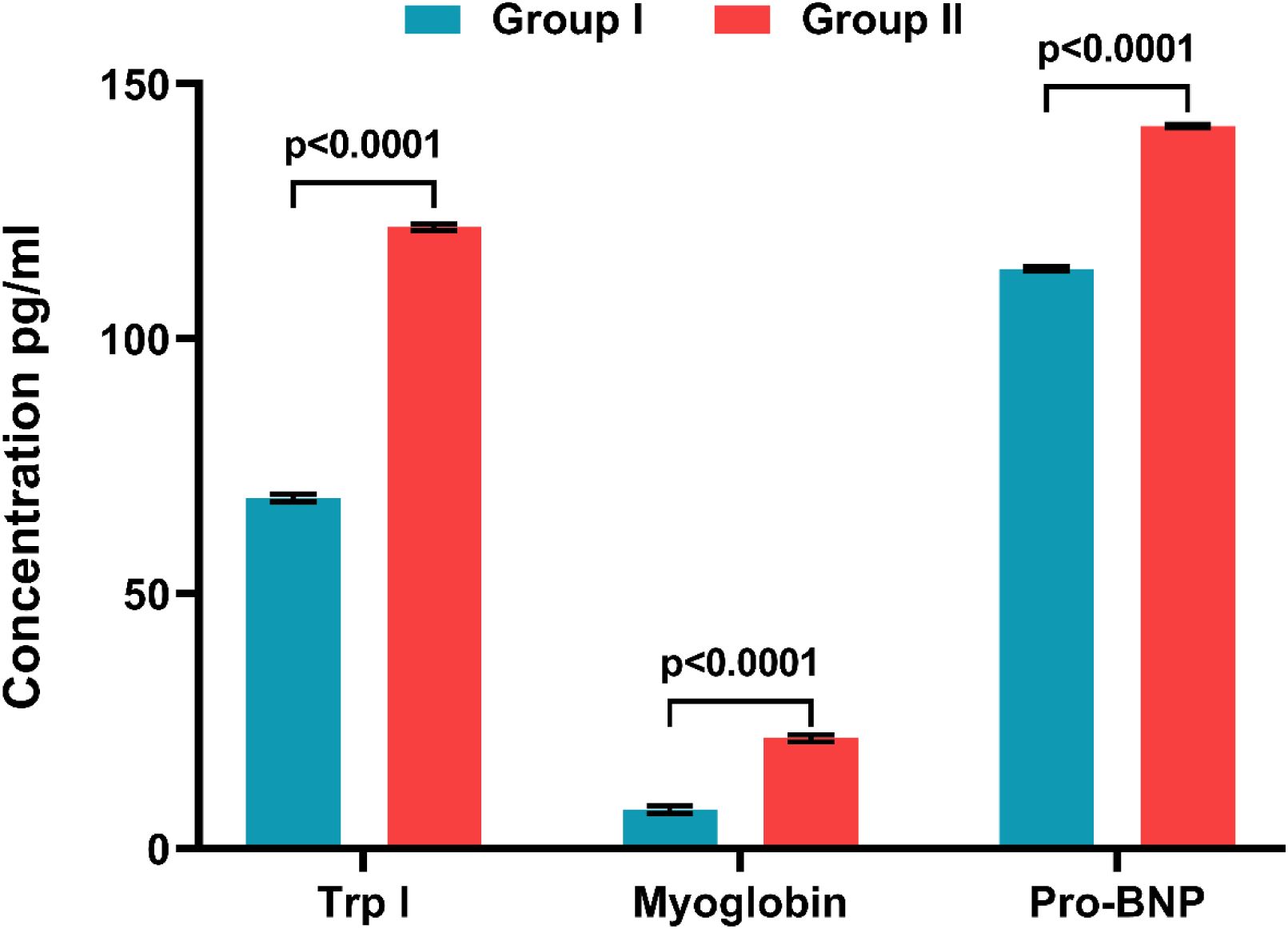
CVD Biomarkers. The graph representing the analysis of cardiac biomarkers expression in group I and group II. The values are expressed as mean ± SE. NS represents non-significant, while the asterisk denotes p ≤ 0.05.

### 3.15. Correlation analysis

Correlations between chronological age and mtDNA copy number, proBNP and methylation status of each CpG site was assessed by Pearson correlation coefficient (r) and corresponding p values were calculated for all parameters. Analyzing the correlations, we observe a weak negative relationship between age and mtDNA/nDNA ratio, suggesting a slight decrease as age increases. Conversely, age exhibits a strong positive correlation with fold change methylation, implying a substantial increase in fold change methylation with advancing age. The correlation between age and proBNP is weakly positive, indicating a modest rise in proBNP levels as individuals age (Figure 14). These interpretations are based on the correlation coefficients, which range from −1 to 1, indicating the strength and direction of linear relationships between variables (Supplementary Table 2).

**Figure 14:**
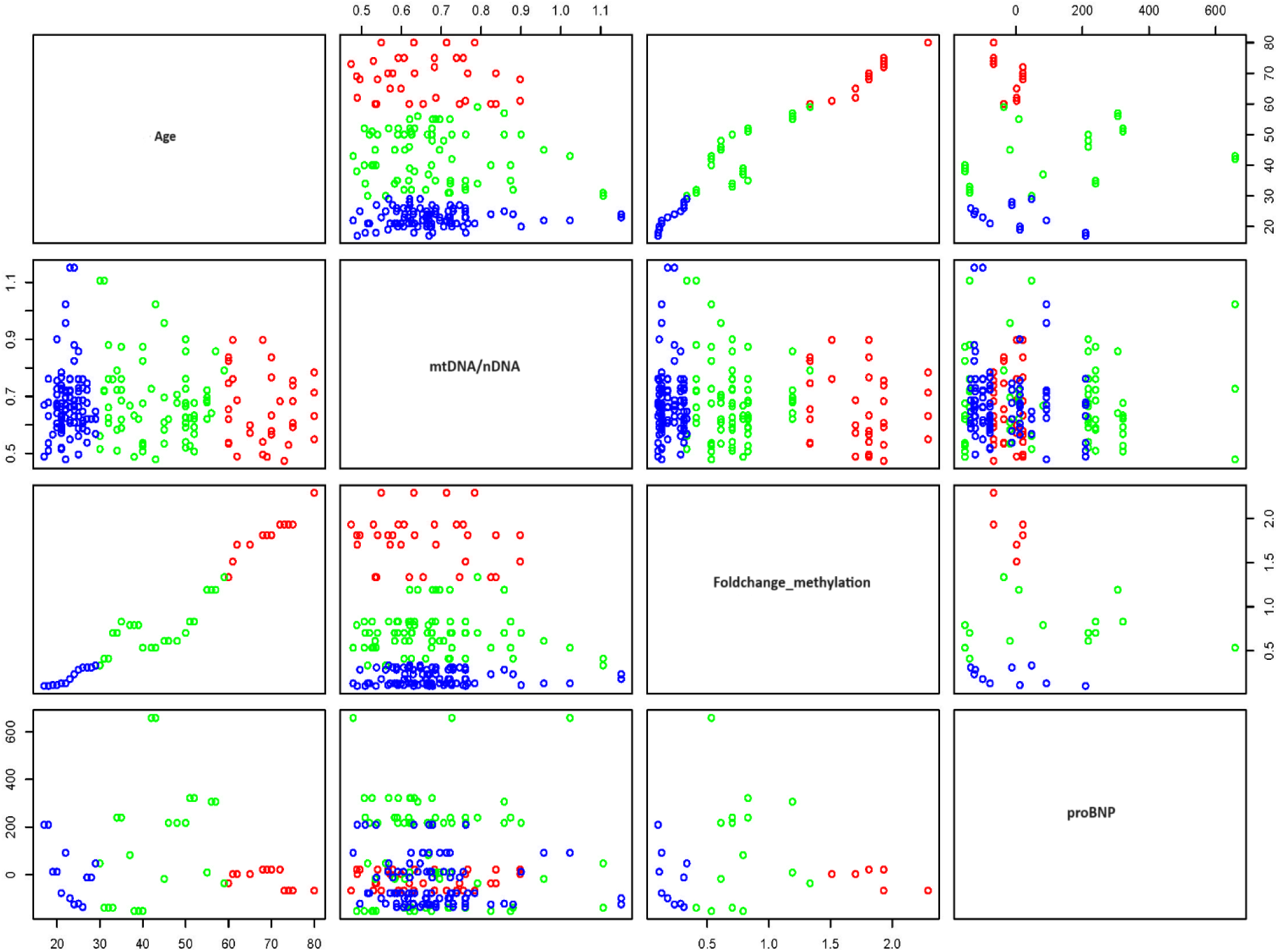
CVD Biomarkers. The graph showing correlations between chronological age and mtDNA copy number, proBNP and methylation status of each CpG site within mtDNA.

## 4. Discussion

This paper reveals that the deterioration of mitochondrial functions could potentially result in the production of reactive oxygen species, influencing the inflammatory status and metabolism of the cell. These alterations may lead to adverse health risks of cardiovascular disorders in elderly people. Using a longitudinal cohort divided among two age groups in India, we found disparities in cardiovascular risk factors linked with mitochondrial dysfunction levels among adults aged 18-65 years and above. It is accompanied by advanced loss of cellular function and systemic deterioration of multiple tissues, leading to an impaired function and increased vulnerability to death. Age-dependent changes distress the receptive capability to maintain normal homeostasis and to tackle stress effectively across a variety of cells including the immune system broadly referred to as immunosenescence. However, development of such effects not only relies on the specific characteristics of the age-related factors like decrease in antioxidant defense mechanisms. Upon sustaining damage, mtDNA, a crucial target for oxidative injury, has the capacity to exacerbate oxidative stress. This amplification occurs through the diminished expression of vital proteins essential for electron transport, initiating a detrimental cycle of free radical generation and dysregulation within the organelles. Consequently, this cascade prompts a response in nuclear DNA damage within the peripheral axis. Despite diverse implications associating cellular responses to DNA damage and repair in cardiac illness with lymphocyte homeostasis, the precise molecular details of this connection remain unexplored. In the lymphocytes of humans, each mitochondrion harbours approximately 10–15 copies of a compact double-stranded circular genome, measuring about 16,569 base pairs. The upkeep of mtDNA is contingent upon the coordinated expression of genes present in both the nucleus and the mitochondria through retrograde signalling. Though a detailed understanding of these pathways is lacking, our initial observations from a cohort of different age individuals suggested the possible association of disturbed epigenetic processes [12]. The development of DNA methylation array technology has facilitated the pinpointing of specific genomic locations for these CpGs. Epigenetic “age estimators” consist of sets of CpGs (referred to as “clock CpGs”) combined with a mathematical algorithm to gauge the age (in years) of a DNA source, such as cells, tissues, or organs. This estimated age, also known as epigenetic age or more precisely DNAm age, not only mirrors chronological age but also reflects the biological age of the DNA source. Due to their precision, DNAm age estimators are commonly denoted as “epigenetic clocks” [24]. Importantly, such disturbances in the vital epigenetic signatures may interfere different mitochondrial functions and cell-fate decisions, are necessary to fine-tune the expression of diverse mitochondrial genes thereby resulting into diseased cellular phenotypes [25]. According to recent studies, controlled mitochondrial-nuclear interaction is crucial for maintaining healthy cellular [8]. Specifically, the metabolic pathways can cause disturbances in mitochondrial machinery and functioning of electron transport chain complexes, premature leakage of electrons and oxidative stress [26]. The mtDNA being in the proximity and lack of protective histones is more vulnerable to such stress and may lead to the formation of guanine oxidation products, a major indicator of oxidative DNA damage. In our study, we observed the higher levels of OGG-1, APE and POLG in older age group, which further confirmed the initiation of BER, upon oxidative damage within the mtDNA of these individuals. The activity of OGG1 is pragmatic herein was almost four folds higher than the APE1. These results were further indicating that APE1 stimulates OGG1 activity, and in the presence of a comparable amount of APE1 the OGG1 can increase up to 5-fold [27]. The pro-survival balancing mechanism can be initiated from the mitochondria and other organelles to maintain the normal cellular homeostasis known as integrated stress response pathway. The ISR molecules coordinates with mitochondrial UPR in an eIF2α dependent mechanism in which the mitochondrial stress associated signals initially activate OMA1 (a protease) to cleave DELE1. After cleavage, DELE1 undergoes accumulation in the cytosol, where it engages with HRI (heme-regulated inhibitor kinase) to facilitate the phosphorylation of eIF2α (eukaryotic initiation factor 2 alpha). Thus, phosphorylated eIF2α assists the translation of specific mRNAs to activate ATF4, which then interacts with ATF3 and CHOP to maintain cellular homeostasis [28,29]. Earlier studies have reported that disturbances in the mitochondrial integrity are closely associated with the induction of ISR [18]. In the current study, the relative quantitative expression of OMA1, DELE1, and HRI genes exhibited modified profiles in the older age risk group. While the increase in HRI was relatively less compared to the fold change observed in OMA1 and DELE1, it was still sufficient to trigger the cascade of Integrated Stress Response (ISR). These findings reinforce our earlier observations, pointing to the induction of ISR attributed to elevated mitochondrial damage associated with aging and disturbed metabolic processes. Mitochondrial stress signals, triggered by free radical injuries, can modulate the expression of genes and proteins involved in mitochondrial-nuclear crosstalk. Accumulating evidence suggests that mitochondrial ROS activates NRF2, recognizing antioxidant response elements (ARE) in specific gene promoter regions and initiating a response. This NRF2-mediated response is facilitated by redox-sensitive thiol(ate) and NF-κB transcription factors, impacting various aspects of mitochondrial physiology and structural integrity [10]. Since aberrant inflammatory responses are seen in both age groups, with higher levels of pro-inflammatory cytokines (IL-6, TNF-α, and IFN-γ) in the older age group, NF-kB activation can boost the release of pro-inflammatory cytokines. Pro-inflammatory cytokine release was noted in both age groups, with higher amounts in the older age group, suggesting a possible connection between aging-related inflammatory processes and NF-κB activation under stressful settings [30]. NRF2, in response to stress, not only counters mitochondrial ROS by upregulating UCP-3 but also influences mitochondrial biogenesis by sustaining PGC-1α levels and promoting purine biosynthesis [31]. Existing literature supports the role of PGC-1α in promoting the expression of genes necessary for mitochondrial biogenesis [32]. In general, the upkeep of the overall shape within cells relies on well-coordinated cycles of fission and fusion as part of mitochondrial biogenesis. Changes in mitochondrial biogenesis, namely in the fission and fusion cycles, are revealed by the study, suggesting that the genome and mitochondrial dynamics may be affected. Group II also exhibits modifications in the expression of fission genes (DRP1, FIS1, and MFF) and fusion genes (OPA1, MFN1, and MFN2). Additionally, there is an observed dysregulation of TFAM and TET genes in the elderly age group, further underscoring the influence of aging on mitochondrial gene expression and epigenetic regulation. In addition, expression profiling of different mitochondrial encoded genes revealed changed levels in group II. The expression of mitochondrial ATPase6, ATPase8, COX1, and ND6 was more disrupted in Group II compared to Group I, accompanied by altered activity of mitochondrial respiratory chain complexes. These data are consistent with earlier research, pointing to a relationship between variations in respiratory chain complex activity among various populations and disruptions of mitochondrial gene expression. Because of its lack of protective histones, limited capacity for repair, and closeness to ROS, mtDNA is especially vulnerable to oxidative damage. Chronic illness risk is elevated and mtDNA depletion can result from mtDNA impairment. The study examines the methylation status of mtDNA regions, revealing significant alterations in mitochondrial methylation profiles associated with age and other environmental factors. This underscores the importance of epigenetic modifications in the context of mitochondrial dysfunction and the higher expression levels of DNA methyltransferase genes (DNMTs) in the group II suggesting a potential link between mitochondrial damage and increased methylation. Furthermore, the analysis of mitochondrial respiratory chain enzymes indicates disturbances in the activities of Complex I-V in the elderly group, providing insights into the functional consequences of mitochondrial dysfunction. Variations in mitomiR expression profiling, which showed differences in mitomiR levels between the two groups and highlighted the function of these short RNAs in controlling the expression of mitochondrial genes and epigenetic modifications in mtDNA [33]. Upon further assessment group II was also found to be associated with more post-translational histone modifications. As errors in epigenetic systems, which control various aspects of chromatin, can affect the transcriptional status of essential genes and cause disease. The detection of CVD biomarkers further strengthens the study’s clinical relevance, with cardiac troponin I show exceptional specificity in identifying cardiac illness in the high-risk group. The various impacts of ageing on mitochondrial dysfunction, repair mechanisms, stress responses, biogenesis, epigenetic regulation, inflammatory processes, and clinical biomarkers are clarified by a thorough discussion that concludes this study. With potential applications for focused treatments and treatment approaches, the findings advance our knowledge of the molecular and cellular pathways underpinning cardiovascular risk in the ageing population.

## 5. Conclusion

In conclusion, this in-depth study thoroughly explores the intricate connection between aging, mitochondrial dysfunction, and cardiovascular diseases, with a specific emphasis on the elderly population. The study underscores the critical role of disruptions in mitochondrial-nuclear crosstalk as critical factors that disturb cellular balance, influence metabolic pathways, and induce oxidative stress. The research delves into the impact of aging on various lymphocyte homeostasis aspects of mitochondrial-mediated epigenetic stress responses, including oxidative stress, inflammatory markers, mtDNA methylation, fission and fusion cycles, expression profiling of mitochondrial genes, alterations in mitochondrial respiratory chain complexes, and shifts in mitomiR levels. These findings emphasize the potential significance of examining mitochondrial dynamics to identify individuals at early risk for cardiovascular events associated with aging. Additionally, future investigations focusing on innovative approaches for detecting mitochondrial dysfunction and epigenetic remodeling could open promising avenues to mitigate the cardiovascular risk, considering the pivotal role mitochondria play in cellular energetics.

## Data Availability

Availability of data and materials: The data that support the findings of this study are available on request from the corresponding author, [PKM].

## Acknowledgements

The authors are thankful to the Department of Health Research (DHR), Ministry of Health & Family Welfare (MoHFW), Government of India, New Delhi for project funding support (R.11013/37/2021-GIA/HR).

## Author’s contribution

PKM: Conceptualization, Supervision, Project administration, Funding acquisition, Writing-original draft preparation, Final approval. NS & PK: Contributed equally, Investigation, Writing-original draft preparation, Final approval. VG, AB, VSR: Investigation, Visualization, Final approval. RT & RKS: Editing, Final approval.

## Ethical approval

The study was approved by the Institutional Human Ethics Committee (IHEC, ICMR-National Institute for Research in Environmental Health, Bhopal, India) and the guidelines of the Indian Council of Medical Research (ICMR), Department of Health Research (DHR), Ministry of Health and Family Welfare (MoHFW), Government of India, were followed.

## Availability of data and materials

The data that support the findings of this study are available on request from the corresponding author, [PKM].

## Abbreviations

AMPK: AMP-activated protein kinase
APE: apurinic/apyrimidinic (AP) endonuclease
ATF-3: cyclic AMP-dependent transcription factor
BER: base excision repair
BMI: body mass index
cGAS-STING: cyclic GMP–AMP synthase stimulator of interferon gene
CHOP: C/EBP homologous protein
CVDs: cardiovascular diseases
DELE1: DAP3 binding cell death enhancer 1
DNMT1: DNA (cytosine-5)-methyltransferase 1
Drp1: dynamin-related protein 1
DSBR: double-strand break repair
eIF2α: eukaryotic initiation factor-2α
Fis1: mitochondrial fission 1 protein
fpg: formamydopyrimidine-DNA glycosylase
HRI: heme-regulated kinase
INF-γ: interferon gamma
Mb: myoglobin
Mff: mitochondrial fission factor
MFN: mitofusins
MMR: mismatch repair
MT-CO1: mitochondrially encoded cytochrome c oxidase
mtDNA: mitochondrial DNA
NADPH: nicotinamide adenine dinucleotide phosphate hydrogen
ND-6: NADH dehydrogenase-6
NF-κB: nuclear factor kappa-light-chain-enhancer of activated B cells
NRF2: nuclear factor erythroid-2-related factor 2
OGG: 8-oxoguanine DNA glycosylase
OMA1: overlapping proteolytic activity with m-AAA protease 1
OPA1: optic atrophy 1
OXPHOS: oxidative phosphorylation
PGC-1α: peroxisome proliferator–activated receptor gamma coactivator-1 alpha
POLG: DNA polymerase subunit gamma
Pro-BNP: pro–b-type natriuretic peptide
ROS: reactive oxygen species
SSBR: single-strand break repair
TET: ten-eleven translocation
TFAM: mitochondrial transcription factor A
TNF-α: tumor necrosis factor-α
TNI: troponin I
UPRmt: unfolded protein response

