## Supplementary Table 1 and Supplementary Table 2 for "Molecular insights into mitoepigenetic stress response signaling in age-associated cardiovascular disease risk"

**SUPPLEMENTARY FILES**

**Table 1. Demographic data of group I (individuals aged 18-38 years) and group II (individuals aged 39-65 years and older).**

| Variables | Group 1 (n=154)  (18-38 years) | Group 2 (n=105)  (39-65 years) |
| --- | --- | --- |
| BMI (kg/m^2^) | 24.3 | 25.04 |
| Age (years)  (Mean ± SD) | 25±5.34 | 57±11 |
| Waist circumference (cm) | 86±5 | 110 ±12 |
| Low blood pressure | 1.7 % | 2.4 % |
| High blood pressure | 2% | 9.3 % |
| High cholesterol (mg/dL) | - | 250±10.52 |
| Troponin I | 68.58±8.34 | 120.76±10.06 |
| Diabetic | - | 5 % |
| COVID-19 history |  | 3.46 % |

Note. All variables present mean ± SE or percent of total subjects with data for each variable.

BMI body mass index, the P value was analysed using chi-squared test.

**Table 2.	Correlation between methylation, mtDNA/nDNA copy number, proBNP and age in all samples.**

| **Pearson coefficient correlation** | | | | |
| --- | --- | --- | --- | --- |
| **Variables** | **Age** | **mtDNA/nDNA** | **Fold change methylation** | **proBNP** |
| **Age** | 1 | -0.07193 | 0.962354 | 0.180244 |
| **mtDNA/nDNA** | -0.07193 | 1 | -0.08207 | -0.02361 |
| **Fold change methylation** | 0.962354 | -0.08207 | 1 | 0.078779 |
| **proBNP** | 0.180244 | -0.02361 | 0.078779 | 1 |
